# Identifying risk factors associated with death among patients with MDR-TB in KwaZulu-Natal, South Africa: an illustration using Weibull parametric model

**DOI:** 10.1101/2022.03.01.22271638

**Authors:** Sizwe Vincent Mbona, Henry Mwambi, Retius Chifurira

## Abstract

**Background:** This study aim was to identify the risk factors associated with multidrug-resistant tuberculosis (MDR-TB) disease. The Weibull model has shown to perform better than the Cox proportional models with respect to the accuracy and efficient of the estimates. Therefore, a Weibull parametric model was employed to identify predictors of death in patients with MDR-TB and the efficiency of the models using current dataset.

**Methods:** Patients diagnosed with MDR-TB were studied in four decentralised sites located in rural areas and one centralised hospital in KwaZulu-Natal, South Africa from July 2008 to July 2012. Patients were followed from the date of MDR-TB diagnosis until death or the last follow-up date.

**Results:** A total of 1 542 patients were included in the analyses: 812 and 730 from the centralised hospital and decentralised sites, respectively. Of the 1 542 enrolled, 15.9% patients died. We found that the hazard of death was significantly higher among patients treated in decentralised sites (aHR) = 1.84, 95% CI = 1.38 – 2.75; SE = 0.81 than that of those who were treated in the centralised hospital. However, the results from the Cox PH model showed an insignificant hazard of death between the decentralised sites and the centralised hospital (aHR = 1.46, 95% CI = 0.69 – 2.36; SE = 0.92). Patients who are between 31 – 40 years of age had increased hazard of death compared to those between 18 – 30 years (aHR = 1.52, 95% CI = 1.04 - 2.23). The hazard of death in female patients was 24% higher compared to male patients (aHR = 1.24, 95% CI = 0.93 - 1.63). Furthermore, patients with previous MDR-TB episodes had an increased hazard of death (aHR = 1.79, 95% CI = 0.23 – 0.62) compared to those with no previous MDR-TB episodes. The hazard of death in HIV negative patients was low compared to those who were HIV positive (aHR = 0.95, 95% CI = 0.57 – 0.77).

**Conclusion:** More health facilities are needed especially in decentralised places and that can help the 2030 World Health Organisation strategy to reduce or end TB infection.

## Background

Multidrug-resistant tuberculosis (MDR-TB) still represent a challenge for clinicians and staff operating in national TB programmes all over the world ^[1-6]^. The World Health Organization (WHO) seeks global evidence on the safety and tolerability of new treatment regimens for drug-resistant tuberculosis, including MDR-TB ^[7]^. Treatment for MDR-TB is challenging for patients, relatives, healthcare providers and health systems ^[8]^.

A number of global studies, including two systematic reviews, have reported higher costs associated with managing MDR-TB patients in hospital ^[9-13]^. South Africa remains one of the highest burdened countries in all three WHO-defined tuberculosis categories, including drug susceptible TB, MDR-TB, and TB/HIV coinfection cases. The previous tuberculosis drug resistance survey done in South Africa during 2001–2002 reported the prevalence of MDR-TB as 1.6% (95% CI 1.1–2.1) in new TB cases and 6.6% (4.9–8.2) in retreatment cases ^[14]^. At that time, the prevalence of TB and HIV was rising, late presentation was common, and tuberculosis-related mortality was high, whereas laboratory testing for drug-resistant tuberculosis was limited. The province of KwaZulu-Natal has amongst the highest prevalence of patients with MDR-TB in South Africa ^[15]^. In settings with limited resources and high prevalence of MDR-TB a decentralized model of care has proven to be effective, and is advisable ^[16]^. In this latter case, only complicated cases are referred to specialized centres or proposed to local/international TB consilia.

Elimination of TB by 2035 will only be possible if countries address the emergence of drug-resistant (DR) strains of *Mycobacterium tuberculosis* effectively. According to the WHO 2018 report, not all DR-TB cases are diagnosed (only 51% of people with bacteriologically confirmed TB were tested for rifampicin resistance (RR) in 2018), and not all DR-TB cases were treated (only one in three of the approximately half a million people who developed MDR/RR-TB in 2018 were treated). DR-TB continues to be an important public health priority ^[17]^, and an estimated 19 million people are latently infected with MDR-TB ^[18]^. The main aim of this study was to employ a Weibull parametric model for better estimates and more flexible than Cox semi-parametric model which is mostly used by many researchers because of its fewer assumptions. The outcome considered here was time from MDR-TB diagnosis until death.

## Methods

### Source of data and description

This was a prospective health systems study including all patients with confirmed diagnosis of MDR-TB, and who commenced treatment between 1 July 2008 and 30 June 2010. Data were sourced from five sites: The Greytown, Manguzi, Murchison, Thulasizwe (Decentralised sites) and King George (Centralised hospital). The data set consists of 1 542 patients, aged 18 years and older, diagnosed with MDR-TB. The target population was defined as all MDR-TB patients diagnosed and treated in the TB centres during the study period. Patients receiving care at more than one site were excluded in order to guarantee the quality of information on MDR-TB treatment episodes. An automatic monitoring method adopted by ^[19]^ sought to eliminate duplicates and correct classification errors of different treatment episodes from the same patient. Inclusion criteria for the comparison study required that patients reside within the catchment area of the site. No data was collected after 1 October 2012 as the study period was from 1 July 2008 to 30 June 2012.

Each participant was examined and followed through regular culture smear sputum tests for MDR-TB outcomes. Conversion of sputum culture from positive to negative was considered a useful early indicator of programme effectiveness, as treatment outcomes were only available 18-24 months after treatment started. Culture conversion was defined as two consecutive negative sputum cultures taken at least one month apart ^[20-22]^. These patients were followed from the date of MDR-TB diagnosis until they die or the last follow-up date.

Medical records were reviewed to collect patient-related demographic, clinical, pharmaceutical and laboratory data. All data, was collected prospectively, prior to knowledge of patient treatment outcomes. Health system data was collected from different components of the health system - laboratory, pharmaceutical and transport services and human resources using existing records and databases, structured questionnaires, observation and interviews. An iterative approach was used which enabled the team to identify new health system data required and develop appropriate data collection methodologies. Over the four-year study period each patient was visited monthly for a day. During each visit data from each health system component was collected, the functioning of the MDR-TB unit observed and informal discussions held with the nurse-in-charge of the MDR-TB unit, the clinician responsible for MDR-TB and the hospital pharmacist. Through a process of ongoing reflection, feedback and discussion with facility and district level staff problems were investigated to determine their origin and cause and possible solutions identified. Field notes detailing the visit and documenting observations and discussions with staff were written up after returning from the site. Notes were also made of concerns, opinions and issues which needed follow up.

The study protocol was approved by the University of KwaZulu-Natal Biomedical Research Ethics Committee (Ref: BF052/09), and by the KwaZulu-Natal Department of Health. Only secondary data, the data routinely collected by health workers for clinical care was used in this study. To protect patient confidentiality and anonymity the data bases were de-identified and access strictly limited. Informed consent was waived by the ethics committee, since all patient data used were previously collected during the course of routine medical care and did not pose any additional risks to the patients.

### Statistical analyses

In this study the multivariable analysis using Weibull distribution in parametric survival model was performed. Survival time was calculated as the time interval between date of MDR-TB diagnosis and date death and date of the last follow-up (for those who did not have an event of interest). The data set was analysed using the Weibull parametric model and Cox proportional hazards model for comparison purposes to show the superiority of the former model. The data set was analysed using STATA version 16 ^[23]^.

### The Cox proportional hazard model

The most well-known model used in survival analysis is the Cox proportional hazards (PH) model ^[24]^. It is a survival analysis regression model, which describes the relation between the event incidence, as expressed by the hazard function and a set of covariates. In brief, if *T* is the survival time, subject to possible right censorship. The Cox PH model is written as

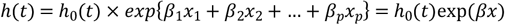

where the hazard function *h*(*t*) is dependent on (or determined by) a set of *p* covariates (*x*_1_, *x*_2_,…, *x*_*p*_), whose impact is measured by the size of the respective coefficients (*β*_1_, *β*_2_,…, *β*_*p*_). The term *h*_0_ is called the baseline hazard, and is the value of the hazard if all the *x*_*i*_ are equal to zero (the quantity exp (0) equals 1). The ‘*t*’ in *h*(*t*) reminds us that the hazard may vary over time. The survival function may be defined in terms of the hazard function by 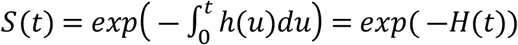, where *H*(*t*) is the cumulative hazard function defined as the area under the hazard function up to time *t*.

In Cox’s PH model, the unknown baseline hazard function *h*_0_ is the non-parametric part and the unknown *β* is the parametric part, which together create a semi-parametric model. Unfortunately, the simplicity of the Cox PH model imposes unrealistic assumptions on the data. Most significantly, the model needs an assumption of independent and identically distributed samples. There are situations, however, where these two assumptions may be found not supported. For example, subjects may be exposed to different risk levels, even after controlling for known risk factors; this is because some relevant covariates are often unavailable to the researcher or even unknown (univariate case).

### The Weibull parametric survival model

The Weibull, which was developed by Waloddi Weibull in 1951, comes originally from engineering issues to analyse the survival data ^[25]^; actually, it has been used to predict the proportion of future failures after observing a failure process at a given point in time ^[26]^. It has a hazard rate which is either increasing, decreasing, or constant ^[27]^. If the hazard rate is constant it will become exponential. Weibull is the only parametric model which has both proportional hazards and an accelerated failure-time representation ^[27]^. In addition, acceptance of Weibull model can be checked via graphical assessment ^[28]^.

The generalization of the exponential distribution to include the shape parameter is the Weibull distribution. The cumulative distribution function of the Weibull distribution is

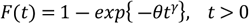

Where *θ* is the shape parameter and *γ* is the scale parameter, and the probability density function of the Weibull distribution is

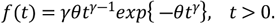

The survival function and hazard function of the Weibull distribution are *S*(*t*) = *exp*{−*θt*^*γ*^} and *h*(*t*) = *γθt*^*γ*−1^respectively. It is easy to see just how flexible the Weibull distribution can be. When *γ* = 1, the Weibull distribution becomes the exponential distribution with *θ* = *λ* and the hazard rate remains constant as time increases, and when *γ* = 2 it is the Rayleigh distribution. For 3 ≤ *γ* ≤ 4, it is close to the normal distribution and when *γ* is large, say *γ* ≥ 10 it is close to the smallest extreme value distribution ^[29]^. When *γ* > 1 the hazard rate increases as time increases, and for *γ* < 1 the hazard rate decreases.

## Results

### Exploratory data analysis

In this analysis study, a total of 1 542 patients diagnosed with MDR-TB were included. A total of 812 (52.7%) patients were treated in the centralised hospital and 730 (47.3%) were treated in the decentralised sites located in rural arears. The mean (± standard deviation) age at the time of diagnosis was 35.7 ± 10.8 years and patients’ age ranged from 18 to 79 years. There were 745 (48.3%) men and 797 (51.7%) women in the study. We further observe that 1 475 (95.7%) had no previous MDR-TB episodes, 64 (4.2%) had one MDR-TB episode and 3 (0.2%) had two or more MDR-TB episodes. The results show that 1 510 (97.9%) patients had pulmonary TB and 32 (2.1%) had extra-pulmonary TB. We observed 864 (56.0%) of the patients who were cured, 245 (15.9%) died, 334 (21.7%) defaulted and 99 (6.4%) were lost in follow-up, during the study period (Table 1).

**Table 1:** Baseline characteristics of patients with MDR-TB enrolled in decentralised and centralised hospitals.

| Factors Categories | Centralised hospital |  | Decentralised sites |  | Total |  | P-value |
| --- | --- | --- | --- | --- | --- | --- | --- |
|  | No | % | No | % | No | % |  |
|  | 812 | 52.7 | 730 | 47.3 | 1 542 | 100 |  |
| Age, mean (SD) | 35.7 (10.8) |  |  |  |  |  |  |
| *Age at diagnosis* |  |  |  |  |  |  |  |
| 18 - 30 | 303 | 19.7 | 245 | 15.9 | 548 | 35.5 | 0.247 |
| 31 - 40 | 292 | 18.9 | 258 | 16.7 | 550 | 35.7 |  |
| 41 - 50 | 145 | 9.4 | 153 | 9.9 | 298 | 19.3 |  |
| 51 or more | 72 | 4.7 | 74 | 4.8 | 146 | 9.5 |  |
| *Gender* |  |  |  |  |  |  |  |
| Male | 399 | 25.9 | 346 | 22.4 | 745 | 48.3 | 0.264 |
| Female | 413 | 26.8 | 384 | 24.9 | 797 | 51.7 |  |
| *Previous MDR-TB episodes* |  |  |  |  |  |  |  |
| No previous MDR-TB episodes | 802 | 52.0 | 673 | 43.6 | 1 475 | 95.7 | <0.001 |
| 1 previous MDR-TB episode | 9 | 0.6 | 55 | 3.6 | 64 | 4.2 |  |
| 2 or more previous MDR-TB episodes | 1 | 0.1 | 2 | 0.1 | 3 | 0.2 |  |
| *Type of TB* |  |  |  |  |  |  |  |
| Pulmonary TB | 804 | 52.1 | 706 | 45.8 | 1 510 | 97.9 | 0.001 |
| Extra-pulmonary TB | 8 | 0.5 | 24 | 1.6 | 32 | 2.1 |  |
| *Comorbidities or other conditions* |  |  |  |  |  |  |  |
| No other diseases or conditions | 780 | 92.5 | 9 | 1.1 | 789 | 93.6 | <0.001 |
| Diabetes | 10 | 1.2 | 10 | 1.2 | 20 | 2.4 |  |
| Epilepsy | 4 | 0.5 | 8 | 1.0 | 12 | 1.4 |  |
| Hearing loss prior to start treatment | 1 | 0.1 | 10 | 1.2 | 11 | 1.3 |  |
| Renal problems | 0 | 0 | 3 | 0.4 | 3 | 0.4 |  |
| Substance abuse | 0 | 0 | 4 | 0.5 | 4 | 0.5 |  |
| Psychiatric problems | 4 | 0.5 | 0 | 0 | 4 | 0.5 |  |
| *HIV status* |  |  |  |  |  |  |  |
| Positive | 576 | 39.1 | 524 | 35.6 | 1 100 | 74.7 | 0.089 |
| Negative | 211 | 14.3 | 162 | 11.0 | 373 | 25.3 |  |
| *TB outcomes* |  |  |  |  |  |  |  |
| Cured | 439 | 28.5 | 425 | 27.5 | 864 | 56.0 | <0.001 |
| Died | 113 | 7.3 | 132 | 8.6 | 245 | 15.9 |  |
| Defaulted | 229 | 14.9 | 105 | 6.8 | 334 | 21.7 |  |
| Lost to follow-up | 31 | 2.0 | 68 | 4.4 | 99 | 6.4 |  |

### Results from the Weibull parametric survival model: multivariable model

Since very few patients had previous MDR-TB episodes and very few had other comorbidities, we categorized the two factors as ‘yes’ or ‘no’ when doing analysis. Results for the univariable analysis showed that study sites, age at diagnosis, gender, previous MDR-TB episodes, and HIV status had a high significant effect on survival time (Table 2).

**Table 2:**
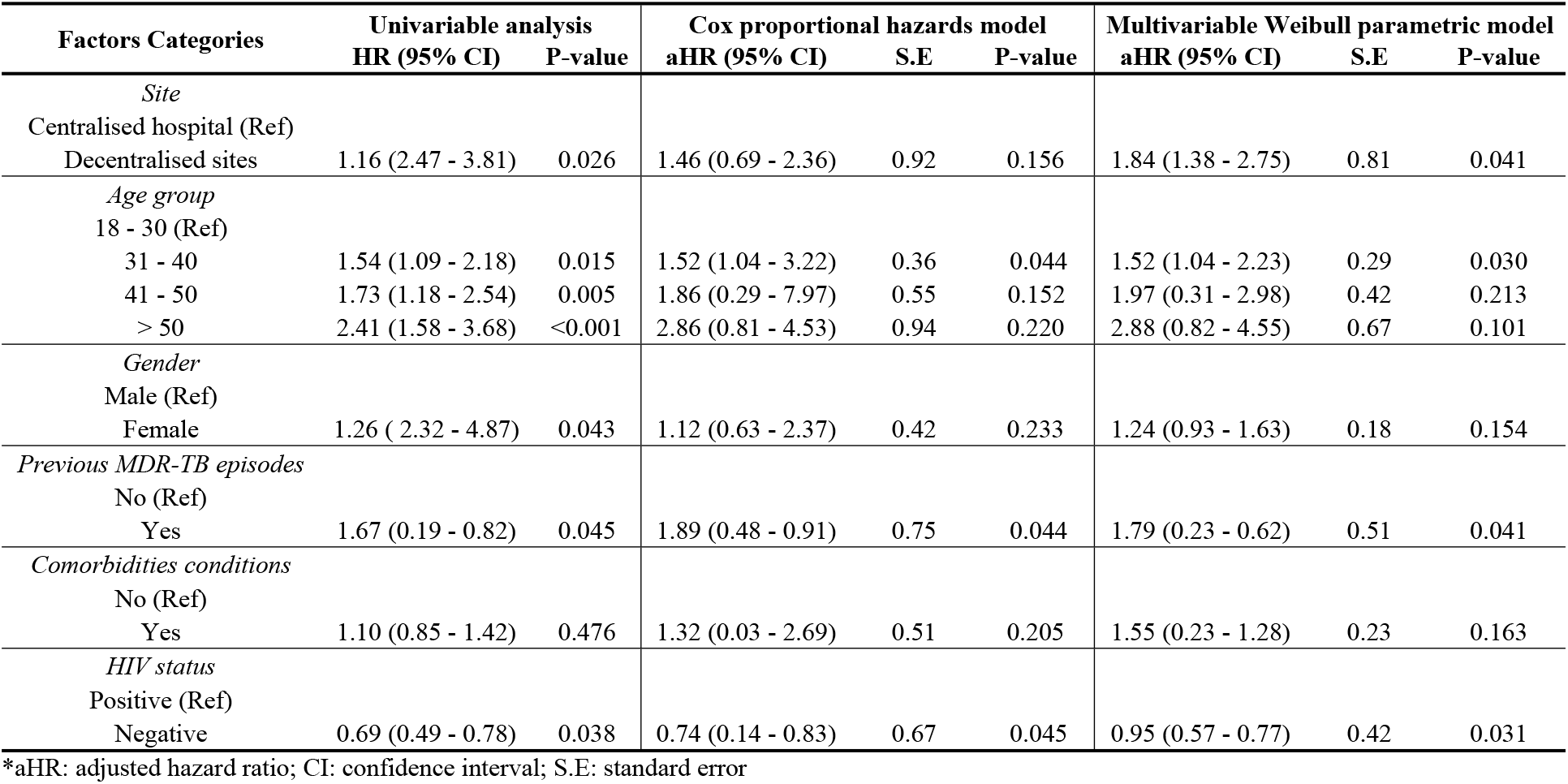
Multivariable Analysis of Weibull Parametric Model

| Factors Categories | Univariable analysis |  | Cox proportional hazards model |  |  | Multivariable Weibull parametric model |  |  |
| --- | --- | --- | --- | --- | --- | --- | --- | --- |
|  | HR (95% CI) | P-value | aHR (95% CI) | S.E | P-value | aHR (95% CI) | S.E | P-value |
| *Site* |  |  |  |  |  |  |  |  |
| Centralised hospital (Ref) |  |  |  |  |  |  |  |  |
| Decentralised sites | 1.16 (2.47 - 3.81) | 0.026 | 1.46 (0.69 - 2.36) | 0.92 | 0.156 | 1.84 (1.38 - 2.75) | 0.81 | 0.041 |
| *Age group* |  |  |  |  |  |  |  |  |
| 18 - 30 (Ref) |  |  |  |  |  |  |  |  |
| 31 - 40 | 1.54 (1.09 - 2.18) | 0.015 | 1.52 (1.04 - 3.22) | 0.36 | 0.044 | 1.52 (1.04 - 2.23) | 0.29 | 0.030 |
| 41 - 50 | 1.73 (1.18 - 2.54) | 0.005 | 1.86 (0.29 - 7.97) | 0.55 | 0.152 | 1.97 (0.31 - 2.98) | 0.42 | 0.213 |
| > 50 | 2.41 (1.58 - 3.68) | <0.001 | 2.86 (0.81 - 4.53) | 0.94 | 0.220 | 2.88 (0.82 - 4.55) | 0.67 | 0.101 |
| *Gender* |  |  |  |  |  |  |  |  |
| Male (Ref) |  |  |  |  |  |  |  |  |
| Female | 1.26 ( 2.32 - 4.87) | 0.043 | 1.12 (0.63 - 2.37) | 0.42 | 0.233 | 1.24 (0.93 - 1.63) | 0.18 | 0.154 |
| *Previous MDR-TB episodes* |  |  |  |  |  |  |  |  |
| No (Ref) |  |  |  |  |  |  |  |  |
| Yes | 1.67 (0.19 - 0.82) | 0.045 | 1.89 (0.48 - 0.91) | 0.75 | 0.044 | 1.79 (0.23 - 0.62) | 0.51 | 0.041 |
| *Comorbidities conditions* |  |  |  |  |  |  |  |  |
| No (Ref) |  |  |  |  |  |  |  |  |
| Yes | 1.10 (0.85 - 1.42) | 0.476 | 1.32 (0.03 - 2.69) | 0.51 | 0.205 | 1.55 (0.23 - 1.28) | 0.23 | 0.163 |
| *HIV status* |  |  |  |  |  |  |  |  |
| Positive (Ref) |  |  |  |  |  |  |  |  |
| Negative | 0.69 (0.49 - 0.78) | 0.038 | 0.74 (0.14 - 0.83) | 0.67 | 0.045 | 0.95 (0.57 - 0.77) | 0.42 | 0.031 |

\*aHR: adjusted hazard ratio; CI: confidence interval; S.E: standard error

The graph of the –ln(-ln(survival probability))) against log of failure time followed a linear like trend which indicated that Weibull model is appropriate for the dataset (Figure 2). Multivariable analysis approach was then performed using study site, age group, gender, previous MDR-TB episodes, comorbidities conditions and HIV status variables. These variables were used regardless of whether the variable was found to be significant or not on the univariate analysis.

We found that the hazard of death was higher among patients treated in decentralised sites (adjusted hazard ratio (aHR) = 1.84, 95% confidence interval (CI) = 1.38 - 2.75) than that of those who were treated in the centralised hospital. This was also supported by the graph in Figure 1. Patients who were between 31 – 40 years of age had increased hazard of death compared to those between 18 – 30 years (aHR = 1.52, 95% CI = 1.04 - 2.23). The hazard of death in female patients was high compared to male patients (aHR = 1.24, 95% CI = 0.93 - 1.63), however this was not significant. Furthermore, patients with previous MDR-TB episodes had an increased hazard of death (aHR = 1.79, 95% CI = 0.23 - 0.62) compared to those with no previous MDR-TB episodes. The result showed that the hazard of death in HIV negative patients was low compared to those who were HIV positive (aHR = 0.95, 95% CI = 0.57 - 0.77).

**Figure 1:**
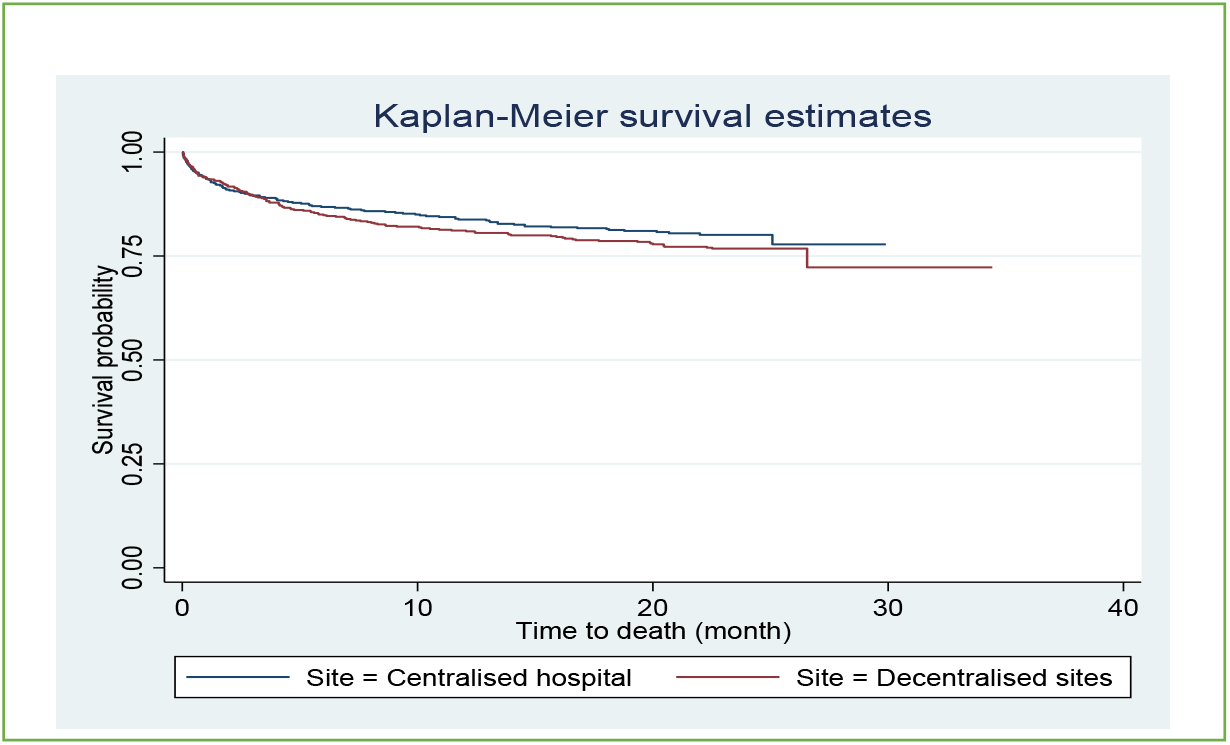
Comparison of sites for time to death in patients treated at a centralised hospital and decentralised sites, 1 July 2008 – 30 June 2012.

**Figure 2:**
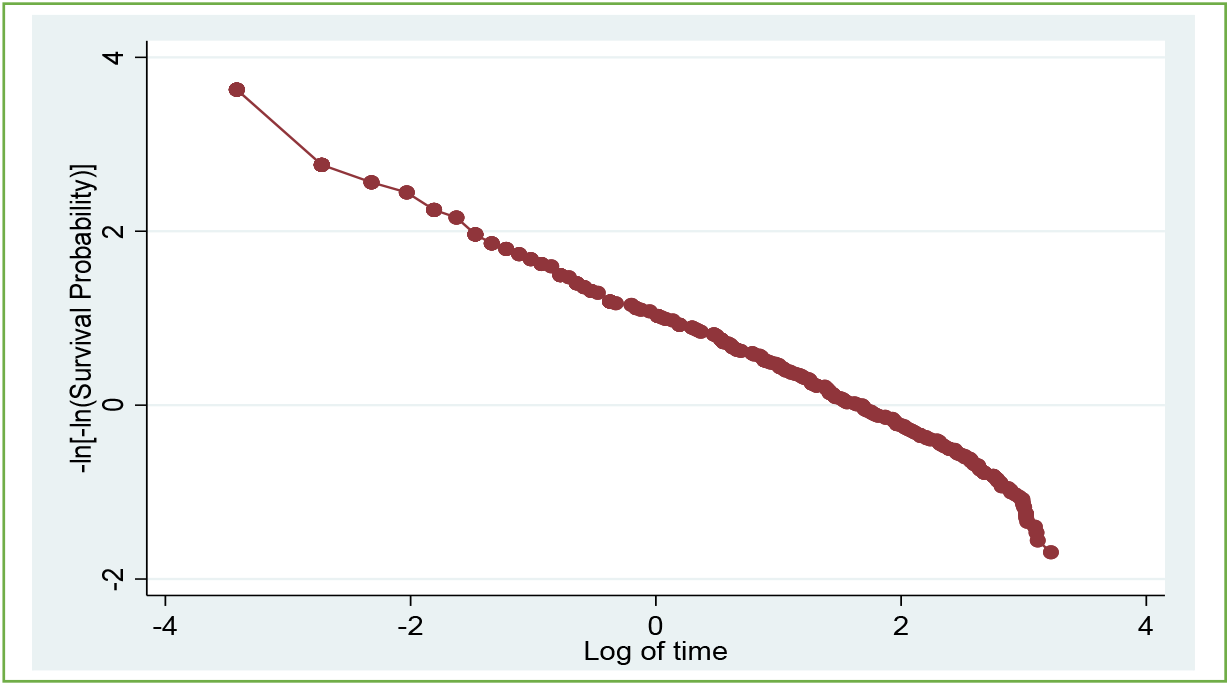
Negative Log of Negative Log Survival Function Estimates

## Discussion

Sub-Saharan Africa is currently seeing a very large change in the major health problems it faces. In this study, the interest was on estimating the survival with multi-explanatory variables, using the Weibull parametric model. Parametric models provide an interpretation based on a particular distribution of the time to event irrespective of proportional hazard assumptions. Most researchers are interested in using the Cox proportional hazard model which in many situations ends up using the last observation of the time-varying covariate. Actually in the case of long-term survival, death rate models are better than Cox model ^[30]^. The standard errors for the estimates in the Cox model were large and in the Weibull parametric model, they were small which makes it a better model for analysis survival time. Furthermore, the fact that Weibull model take into account time varying covariates, allow researcher some flexibility. Weibull model was advantageous for modelling multivariable as it has a hazard rate which is either increasing, decreasing, or constant ^[27]^.

The main aim of this study was to identify risk factors of death among patients with MDR-TB disease. Drawing on our findings based on Multivariable Weibull model, site, age at diagnosis, previous MDR-TB episodes and HIV status were significant. The results of this showed that, in overall, 56% of the patients were cured of MDR-TB, 15.9% died during the follow-up, 21.7% were defaulted and 6.4% were lost in follow-up. Results from the Cox model showed no statistical significant difference between the centralised hospital and decentralised sites in terms of dying over time, with patients in decentralised sites having higher hazard of death than those in the centralised hospital. Results from Weibull parametric model indicated that the death rate was significantly different between the centralised hospital and the decentralised sites and the standard errors were small compared to those obtained in Cox model. Most risk factors that was identified by Weibull model are those identified but Cox model but Weibull model was superior because produced parameter estimates with small standard errors. Carroll’s study indicated that in the analysis of survival data Weibull model can provide a useful, parametric alternative to Cox’s regression modeling ^[31]^.

There are some limitations to this study. we did not have any information about the socioeconomic factors of patients but interest may be done either by investigating the effect of these factors. In addition, our study was carried out based on data collected in KwaZulu-Natal province, South Africa and therefore the findings could not be generated.

## Conclusion

The Weibull model enables the assessment of the effect of factors while taking into account the distribution of the data. It is the best option for analyzing lifetime data if the distributional assumptions can be met and the shape parameter is known. However, when the shape parameter is unknown, the Cox proportional hazards model is a good alternative. It requires fewer assumptions than the parametric Weibull model and provides comparable mean square errors of the estimates of PH-slope. We conclude that even in resource-limited settings and in the presence of HIV co-infection, community-based care is more effective as care in either a centralized or decentralized hospital setting for patients who do not require hospitalization and that could decrease the number of death due to tuberculosis as number of infection continue to increase.

## Data Availability

All relevant data are within the manuscript and its Supporting Information files.

## Abbreviations

aHR: Adjusted hazard ratio
CI: confidence interval
S.E: standard error
SD: Standard deviation
HIV: Human immunodeficiency virus
TB: Tuberculosis
DR: Drug-resistant
MDR-TB: Multidrug-resistant tuberculosis

## Declarations

### Consent for publication

Not applicable

### Availability of data and materials

Data will be made available upon request but will be controlled.

### Authors’ contributions

All the authors made contribution to the study. SVM planned the study and wrote the initial draft of the article and did the analysis. HW and RC assisted with data analysis and interpretation. SVM did the revisions to the paper assisted by HW and RC. All authors approved submission of this article. The author(s) read and approved the final manuscript.

## Acknowledgements

We thank Dr Marian Loveday and Prof Glenda Matthews for allowing us to use their dataset. We thank all facility level managers, doctors, nurses and data capturers at the study sites for their assistance. Thank you all very much!

## Authors’ information

**Mr Sizwe Vincent Mbona:** Durban University of Technology, Statistics Department, 41-43 ML Sultan Road, Durban, 4001.

**Prof. Henry Mwambi:** University of KwaZulu-Natal, School of Mathematics, Statistics and Computer Science, King Edward Avenue, Pietermaritzburg, 3209.

**Dr Retius Chifurira:** University of KwaZulu-Natal, School of Mathematics, Statistics and Computer Science, University Road, Westville, 4000.

## References

[1] Akkerman O, Aleksa A, Alffenaar JW, Al-Marzouqi NH, Arias-Guillén M, Belilovski E, Bernal E, Boeree MJ, Borisov SE, Bruchfeld J, Loidi JC. Surveillance of adverse events in the treatment of drug-resistant tuberculosis: A global feasibility study. International Journal of Infectious Diseases. 2019; 83:72–6.

[2] Borisov S, Danila E, Maryandyshev A, Dalcolmo M, Miliauskas S, Kuksa L, Manga S, Skrahina A, Diktanas S, Codecasa LR, Aleksa A. Surveillance of adverse events in the treatment of drug-resistant tuberculosis: first global report. European Respiratory Journal. 2019; 54(6).

[3] Borisov SE, Dheda K, Enwerem M, Leyet RR, D’Ambrosio L, Centis R, Sotgiu G, Tiberi S, Alffenaar JW, Maryandyshev A, Belilovski E. Effectiveness and safety of bedaquiline-containing regimens in the treatment of MDR-and XDR-TB: a multicentre study. European Respiratory Journal. 2017; 49(5).

[4] Lange C, Aarnoutse RE, Alffenaar JW, Bothamley G, Brinkmann F, Costa J, Chesov D, Van Crevel R, Dedicoat M, Dominguez J, Duarte R. Management of patients with multidrug-resistant tuberculosis. The international journal of tuberculosis and lung disease. 2019; 23(6):645–62.

[5] Nahid P, Mase SR, Migliori GB, Sotgiu G, Bothamley GH, Brozek JL, Cattamanchi A, Cegielski JP, Chen L, Daley CL, Dalton TL. Treatment of drug-resistant tuberculosis. An official ATS/CDC/ERS/IDSA clinical practice guideline. American journal of respiratory and critical care medicine. 2019; 200(10): 93–142.

[6] Migliori GB, Global Tuberculosis Network (GTN). Evolution of programmatic definitions used in tuberculosis prevention and care. Clinical Infectious Diseases. 2019; 68(10):1787–9.

[7] Halleux, Christine M., Dennis Falzon, Corinne Merle, Ernesto Jaramillo, Fuad Mirzayev, Piero Olliaro, and Karin Weyer. “The World Health Organization global aDSM database: generating evidence on the safety of new treatment regimens for drug-resistant tuberculosis.” 2018: 1701643.

[8] Lange C, Abubakar I, Alffenaar JW, Bothamley G, Caminero JA, Carvalho AC, Chang KC, Codecasa L, Correia A, Crudu V, Davies P. Management of patients with multidrug-resistant/extensively drug-resistant tuberculosis in Europe: a TBNET consensus statement. 2014: 23–63.

[9] Fitzpatrick C, Floyd K. A systematic review of the cost and cost effectiveness of treatment for multidrug-resistant tuberculosis. Pharmacoeconomics. 2012; 30(1):63–80.

[10] Bassili A, Fitzpatrick C, Qadeer E, Fatima R, Floyd K, Jaramillo E. A systematic review of the effectiveness of hospital-and ambulatory-based management of multidrug-resistant tuberculosis. The American Journal of Tropical Medicine and Hygiene. 2013; 89(2):271–80.

[11] Floyd K, Hutubessy R, Kliiman K, Centis R, Khurieva N, Jakobowiak W, Danilovits M, Peremitin G, Keshavjee S, Migliori GB. Cost and cost-effectiveness of multidrug-resistant tuberculosis treatment in Estonia and Russia. European Respiratory Journal. 2012; 40(1):133–42.

[12] Schnippel K, Rosen S, Shearer K, Martinson N, Long L, Sanne I, Variava E. Costs of inpatient treatment for multi-drug-resistant tuberculosis in S outh A frica. Tropical Medicine & International Health. 2013; 18(1):109–16.

[13] Cox H, Ramma L, Wilkinson L, Azevedo V, Sinanovic E. Cost per patient of treatment for rifampicin-resistant tuberculosis in a community-based programme in Khayelitsha, South Africa. Tropical Medicine & International Health. 2015; 20(10):1337–45.

[14] Weyer K, Brand J, Lancaster J, Levin J, Van der Walt M. Determinants of multidrug-resistant tuberculosis in South Africa: results from a national survey. South African Medical Journal. 2002; 92(3).

[15] Ismail NA, Mvusi L, Nanoo A, Dreyer A, Omar SV, Babatunde S, Molebatsi T, Van der Walt M, Adelekan A, Deyde V, Ihekweazu C. Prevalence of drug-resistant tuberculosis and imputed burden in South Africa: a national and sub-national cross-sectional survey. The Lancet Infectious Diseases. 2018; 18(7):779–87.

[16] Loveday M, Wallengren K, Reddy T, Besada D, Brust JC, Voce A, Desai H, Ngozo J, Radebe Z, Master I, Padayatchi N. MDR-TB patients in KwaZulu-Natal, South Africa: Cost-effectiveness of 5 models of care. PloS one. 2018; 13(4): 0196003.

[17] World Health Organization. WHO consolidated guidelines on drug-resistant tuberculosis treatment. World Health Organization; 2019.

[18] Knight GM, McQuaid CF, Dodd PJ, Houben RM. Global burden of latent multidrug-resistant tuberculosis: trends and estimates based on mathematical modelling. The Lancet Infectious Diseases. 2019; 19(8):903–12.

[19] Bierrenbach AL, Pinto de Oliveira G, Codenotti S, Gomes AB, Stevens AP. Duplicates and misclassification of tuberculosis notification records in Brazil, 2001–2007. The International Journal of Tuberculosis and Lung Disease. 2010; 14(5):593–9.

[20] Frette C, Jacob MP, Kauffmann F, Mitchison DA. Assessment of new sterilizing drugs for treating pulmonary tuberculosis by culture at 2 months.[correspondence]. Am Rev Respir Dis. 1993;147:1062–3.

[21] Lienhardt C, Davies G. Methodological issues in the design of clinical trials for the treatment of multidrug-resistant tuberculosis: challenges and opportunities [State of the art series. Drug-resistant tuberculosis. edited by CY. Chiang. Number 5 in the series]. The International Journal of Tuberculosis and Lung Disease. 2010; 14(5):528–37.

[22] Laserson KF, Thorpe LE, Leimane V, Weyer K, Mitnick CD, Riekstina V, Zarovska E, Rich ML, Fraser HS, Alarcon E, Cegielski JP. Speaking the same language: treatment outcome definitions for multidrug-resistant tuberculosis. The International Journal of Tuberculosis and Lung Disease. 2005; 9(6):640–5.

[23] StataCorp LP. Stata statistical software. College Station TX. 2019.

[24] Cox DR. Regression models and life-tables. Journal of the Royal Statistical Society: Series B (Methodological). 1972 Jan;34(2):187–202.

[25] Weibull W. Wide applicability. Journal of applied mechanics. 1951; 103(730):293–7.

[26] Nelson W. Weibull prediction of a future number of failures. Quality and Reliability Engineering International. 2000; 16(1):23–6.

[27] Klein JP, Moeschberger ML. Survival Analysis: Techniques for Truncated and Censored Data. 2003.

[28] Kleinbaum DG, Klein M. Survival Analysis: A self-learning text, 2005. New York, NY: Spring Science+ Business Media. 2011.

[29] Nelson W. Applied Life Data Analysis, New York: JohnWiley. NelsonApplied Life Data Analysis. 1982.

[30] Rahimzadeh M, Baghestani AR, Gohari MR, Pourhoseingholi MA. Estimation of the cure rate in Iranian breast cancer patients. Asian Pacific journal of cancer prevention. 2014; 15(12): 4839–42.

[31] Carroll KJ. On the use and utility of the Weibull model in the analysis of survival data. Controlled clinical trials. 2003; 24(6):682–701.

